# Cost-Effectiveness of Coronary Artery Bypass Grafting (CABG) versus Percutaneous Coronary Intervention (PCI) as an add-on strategy to Optimal Medical Therapy (OMT) in Severe Ischemic Cardiomyopathy

**DOI:** 10.1101/2024.06.07.24308637

**Authors:** Shumail Fatima, Gavin W. Hickey, Matthew E. Harinstein, John J. Pacella, Ibrahim Sultan, Kenneth J. Smith

## Abstract

1.

**Background:** Revascularization through coronary artery bypass grafting (CABG) or percutaneous coronary intervention (PCI) as an add-on therapy to optimal medical therapy (OMT) is routinely used in patients with severe ischemic cardiomyopathy (ICM) with ejection fraction (EF) ≤ 35% to improve cardiovascular outcomes with limited available data about their relative costs and efficacies. We performed a cost-effectiveness analysis to illustrate the most economically favorable strategy in this population.

**Methods:** A Markov model simulated a cohort with severe ICM (EF ≤ 35%) and evaluated strategies of CABG+OMT, PCI+OMT and OMT alone. Model inputs were obtained from STICHES and REVIVED clinical trials and their subsequent cost-effectiveness analyses. Cohorts were followed monthly, with mortality and major adverse cardiac events (MACE; a composite of heart failure hospitalization, myocardial infarction, revascularization and arrythmias) modeled as lifetime disutilities, taking the US health system perspective, and discounting 3%/year over lifetime horizon. Outcome measures were lifetime medical costs (2019 US$), quality-adjusted-life-years (QALYs), and incremental cost-effectiveness ratios (ICERs).

**Results:** OMT alone was the least costly strategy at $107,780 and yielded 5.29 QALYs. PCI+OMT yielded 4.87 QALYs and cost $121,368, while the CABG+OMT strategy resulted in 7.01 QALYs and cost $160,124 or $38,755 per QALY gained compared to PCI+OMT with an ICER of $18,130 per QALY gained over lifetime horizon. Thus, CABG+OMT was preferred at a $100,000/QALY gained threshold, a commonly cited US benchmark. In a probabilistic sensitivity analysis, CABG+OMT was the preferred strategy in 69%, 82% and 85%% of the model iterations at $50,000, $100,000, and $150,000 per QALY gained willingness-to-pay (WTP) thresholds respectively.

**Conclusions:** CABG+OMT is the most cost-effective strategy in patients with severe ICM as compared with PCI+OMT or OMT only strategies at current benchmarks for value in the United States.

**WHAT IS KNOWN:**

- Revascularization though coronary artery bypass grafting (CABG) or percutaneous coronary intervention (PCI) are routinely used with optimal medical therapy to treat severe ischemic cardiomyopathy (ICM).
- The STICH (Surgical Treatment for Ischemic Heart Failure) trial demonstrated survival and economic superiority of CABG over OMT whereas the REVIVED (Revascularization for Ischemic Ventricular Dysfunction) trial showed no difference in clinical or economic outcomes between PCI and OMT groups in patients with ICM.
- Currently, no large-scale clinical trials have directly compared the health and economic outcomes of CABG versus PCI as adjunctive therapies to OMT in the ICM population. Consequently, their comparative cost-effectiveness remains unknown.

**WHAT THE STUDY ADDS:**

- This study presents the first cost-effectiveness analysis comparing CABG+OMT versus PCI+OMT strategies in patients with severe ICM, addressing a significant knowledge gap.
- Results indicate that despite being more invasive and higher initial costs, CABG+OMT approach yields superior health outcomes and offers a financial advantage over the PCI+OMT strategy.
- This study identifies CABG+OMT as the most cost-effective strategy over PCI+OMT in severe ICM, thereby aiding decision-making for physicians and policymakers.

## 2. INTRODUCTION

With the prevalence of ischemic heart disease-induced heart failure (HF) reaching epidemic proportions, the augmentation of prognosis-modifying medical therapies with revascularization strategies has become an important consideration to mitigate the associated excess mortality and morbidity^1–4^. By 2030, more than 8 million people in US are projected to suffer from HF and cumulative costs associated with HF care would increase to $70 billion annually^5^. Ischemic cardiomyopathy (ICM) accounts for 60% of HF cases in US annually, underscoring its significant role in the etiology of this condition^6^. Although the introduction of optimal medical therapy (OMT) and preventive device therapy has marked a significant advancement in the management of ICM and heart failure with reduced ejection fraction (HFrEF), patients with severe ventricular dysfunction, as characterized by left ventricular ejection fraction (LVEF) of 35% or lower, remain at high risk for adverse events due to recurrent ischemic insults^7,8^. Revascularization holds particular promise for this patient population, as it offers to limit and reverse the ischemia-driven progression of ventricular damage with ultimate goal to prevent the onset of a decompensated HF state, marked by diminished quality of life, extensive healthcare utilization, and ultimately, premature death^9–11^.

Despite their conceptual similarity, not all revascularization strategies confer equivalent clinical benefits to ICM patients. The STICH Randomized Clinical Trial (Surgical Treatment for Ischemic Heart Failure) showed that supplementing OMT with coronary artery bypass grafting (CABG) significantly improved survival at the 10-year follow-up compared to OMT alone in severe ICM with LVEF ≤35%^12^. Conversely, the REVIVED Randomized Clinical Trial (Revascularization for Ischemic Ventricular Dysfunction) found no significant difference in event-free survival after 41 months when percutaneous coronary intervention (PCI) was added to OMT, as opposed to OMT alone in similar patient population, despite PCI arguably being less invasive and less costly intervention compared with CABG^13^. Beyond these differences in clinical outcomes, the STICH trial identified the CABG+OMT strategy as cost-effective, whereas the REVIVED trial did not recognize PCI+OMT as cost-effective in comparison to OMT alone^14,15^. Despite the demonstrated clinical and economic superiority of CABG over PCI as an adjunct revascularization strategy to OMT in separate trials, a direct and prospective comparison of clinical and financial outcomes between CABG and PCI in severe ICM is yet to be performed.

To overcome this knowledge gap, we are presenting a long-term cost-effectiveness analysis comparing CABG and PCI strategies as add-on therapies to OMT in the context of severe ICM based on trial-level clinical and medical utilization data from STICH and REVIVED clinical trials from US healthcare system perspective.

## 3. METHODS

### Economic model overview

We constructed a Markov state-transition cohort model to estimate the lifetime benefits and costs associated with CABG+OMT, PCI+OMT, and OMT strategies from the perspective of the US healthcare system (Figure S1). This model included patients diagnosed with severe ICM, characterized by LVEF of 35% or lower, following revascularization through CABG or PCI, or solely managed with OMT. Within each treatment strategy, patients could experience death, a major adverse cardiac event (MACE)—a composite of heart failure hospitalization (HFH), myocardial infarction (MI), revascularization (in case of CABG and PCI strategies: post-index-procedure revascularization) and arrhythmias—or remain free from death and MACE in post-intervention health state. These outcomes were evaluated over monthly transition periods. The probabilities of death and MACE for the CABG+OMT, PCI+OMT, and OMT strategies were obtained from the publicly available STICH and REVIVED clinical trials data. Similarly, the costs, disutilities, quality-adjusted life years (QALYs) and life years (LYs) accrued by simulated cohort for these strategies were extracted from the prospectively collected medical resource use and quality of life (QoL) data of the STICH and REVIVED trials. Model input uncertainties were assessed through 1-way sensitivity analyses whereas model results stability was evaluated in probabilistic sensitivity analysis. The model’s outcomes, including costs, QALYs, LYs, and the incremental cost-effectiveness ratio (ICER), were presented based on an intention-to-treat analysis, in accordance with the trials data. As deidentified and publicly available data was used, therefore, institutional review board approval is not required.

### Trial Design and Patient Population

The STICH trial randomized 1212 participants aged 18 years or older with coronary artery disease amendable to CABG and LVEF of 35% or lower to one of the two groups: CABG+OMT group (610 patients) or OMT only group (602 patients)^12^. The median age of the cohort was 60 years, females constituted 12% of the sample and ethnic or racial minorities represented 36%. A history of MI was present in 76% of participants, 69% had significant disease in left main or proximal left anterior descending coronary arteries whereas 36% were diagnosed with triple vessel disease. 54% of the participants had an EF of 28% or less, and 37% were categorized as having New York Heart Association (NYHA) class III or IV heart failure. The follow-up period for this trial extended to a median of 9.8 years. All-cause mortality was the primary outcome of interest.

In the REVIVED-BCIS2 trial, 700 patients with extensive coronary artery disease with LVEF of 35% or less and demonstrable myocardial viability amendable to PCI were randomized to either a strategy of PCI+OMT (347 patients) or only OMT (353 patients)^13^. The median age of the participants was 69 years, 12% were females and only 12% belonged to ethnic or racial minority groups. Similar to STICH, 38% of the participants had triple-vessel disease. However, the percentage of participants with prior history of MI (50%), left main coronary artery disease (14%) and EF of 29% or lower (36%) was lower than that of STICH cohort. Similarly, only 26% had NYHA class III or IV heart failure as opposed to 37% in STICH. The median duration of follow-up was 41 months. The primary outcome was a composite of death from any cause or HFH, whereas the secondary outcomes included the individual components of the primary outcome.

### Probabilities

All-cause mortality data for CABG+OMT group was obtained from the STICH trial, where 58.9% of participants randomized to CABG and 66.1% randomized to medical therapy alone group had experienced death at end of 9.8 years of follow-up^12^. In parallel, all-cause mortality data for the PCI+OMT group was derived from the REVIVED trial indicating 31.7% of patients in the PCI arm and 32.6% in the medical therapy alone arm died during 41-month follow-up period^13^. Given the availability of two distinct mortality probabilities for the medical therapy alone group from both these trials, an average was utilized as the base case for the OMT strategy in the model and the individual probabilities from both the trials served as the upper and lower bounds of the range in the sensitivity analysis.

MACE was defined as a composite outcome for each strategy, including HFH, MI, revascularization, and arrhythmias. These events were selected as components of MACE due to their high prevalence among ICM complications^16–19^. Despite STICH and REVIVED trials reporting these MACE components for both intervention and medical therapy groups, discerning mutually exclusive events within participants was unfeasible due to lack of access to patient-level data. Consequently, assumptions were necessary to approximate the average probability of MACE in each strategy. Under the no overlap assumption (all events considered unique and mutually exclusive), the cumulative probability of MACE was calculated by aggregating individual component probabilities. Conversely, with the maximum overlap assumption (each event presumed to affect every participant until reaching sample size limitations), the probability equaled that of the most frequent event. Given the tendency of the maximum overlap assumption to underestimate and no overlap to overestimate MACE probability, an average of these probabilities was utilized for the base-case scenario for CABG+OMT and PCI+OMT strategies, while individual values served as lower and upper bounds respectively in sensitivity analysis. Following the same methodology, the OMT strategy’s MACE probability was derived as a mean of averages from both assumptions, mirroring the approach taken for all-cause mortality. This step was performed as both trials have reported MACE events for their respective medical therapy alone groups. Finally, all the probabilities were adjusted for monthly rates in the model (Table S1).

### Health-Related Quality of Life

ED-5D response-based utility index values and MACE-related disutilities were incorporated for CABG+OMT, PCI+OMT and OMT strategies as a quality of life (QoL) measure. The STICH trial employed EQ-5D 3-level instrument to capture QoL data at 4, 12, 24, 36 months and translated the responses to a weighted utility index with values ranging from 0 (death) to 1 (perfect health)^14^. We used utility index value at 36 months for our base-case for the CABG+OMT group and assumed that this utility value persisted beyond the trial duration, with reductions due to post-CABG MACE incorporated as disutilities in the model. We varied the base-case utility values over a plausible range encapsulating the utility values obtained at prior timepoints in the sensitivity analysis.

For the PCI+OMT strategy, the utility index was obtained from the REVIVED trial, which evaluated QoL data using EQ-5D-5L and KCCQ questionnaires at baseline, 6 months, 12 months, and annually up to 8 years post-randomization^15^. The EQ-5D-5L utility index value at 36 months was adopted as the base case for PCI+OMT group to align with the STICH trial’s utility values timepoint for the CABG+OMT group. We also assumed that this utility value would persist beyond the trial duration, albeit with adjustments for disutilities resulting from post-PCI MACE. We also varied the utility values over a range that included the values obtained during 8-years in the sensitivity analysis.

For the OMT strategy, because both the STICH and REVIVED trials measured utility indices for their medical therapy only groups, a weighted average was employed for the base-case scenario, with the respective trial values varied over the reported ranges in sensitivity analysis. Analogous to the CABG+OMT and PCI+OMT groups, the utility index for the OMT strategy was adjusted to account for disutilities associated with MACE.

The disutilities for individual MACE components constituted a weighted aggregate of disutility values associated with HFH, MI, revascularization and arrythmias encountered in each strategy. Because disutility values associated with these events in the STICH and REVIVED trials were not reported, estimations were adopted from published literature. The EQ-5D utility value for HFH was obtained from ACSEND-HF post-hoc analysis, while a systematic review provided the EQ-5D utility value for MI^20,21^. Due to a paucity of data about the disutility values for lethal arrhythmias (ventricular tachycardia and ventricular fibrillation etc.), disutility estimates for atrial fibrillation from the CABANA trial were assumed for arrythmias, albeit with an acknowledgment that the disutility values for lethal arrhythmias may be substantially higher^22^. Finally, these values were multiplied with the baseline post-intervention utility values to obtain chronic health state corrected utilities. The average duration of MACE, encompassing length of hospitalization for each unique MACE component was posited at 5 days for base-case based on the published data, and was varied between 3-7 days in sensitivity analysis^23^ (Table S1).

### Cost Estimations

Costs were based on prospective assessments of medical resource utilization during STICH and REVIVED trials durations, including expenses of initial procedures, medications, follow-up appointments, rehospitalizations, and follow-up procedures in both inpatient and outpatient settings^14,15^. STICH quantified these expenses in 2019 US dollars, whereas REVIVED reported in British Pounds in their published cost-effectiveness analyses. As the current analysis is conducted from US healthcare perspective, we opted to use the costs reported by STICH as reference for this analysis and calculated costs for PCI strategy based on REVIVED medical resource use data. STICH derived hospital-based service costs from the Premier Healthcare Database for 2016 while outpatient management costs, including office visits and medications, were sourced from the Medical Expenditure Panel Survey Household component from 2002 to 2011^24,25^. STICH estimated physician costs from the professional fee ratios reported for patients with HF^26^.

Both CABG+OMT and PCI+OMT strategies accrued the upfront index-procedure costs in addition to medications costs as well as office visit costs. One-time cost for implantable cardioversion defibrillator (ICD) was also applied across all strategies based on American College of Cardiology recommendation for ICD in patients with ischemic heart disease with an EF ≤ 35% (Class IA)^27^. Afterwards, CABG+OMT, PCI+OMT and OMT groups were assumed to have equivalent annual medication and outpatient office visit costs in the absence of mortality and MACE. For strategy-specific MACE costs, weighted costs of hospitalization due to HF, MI, revascularization though CABG or PCI and arrythmias were also included. All future costs and benefits were discounted at 3% per year (Table S1).

### Cost-effectiveness Analysis

For the cost-effectiveness analysis, we calculated ICERs as the difference in mean lifetime costs divided by the difference in mean quality-adjusted life years across CABG+OMT, PCI+OMT and OMT strategies. To illustrate the influence of key model input in our base-case analysis on the model outcome estimates, we also performed a range of univariable sensitivity analyses (1-way deterministic analysis) by varying a single input parameter at a time and observing the change in ICER. This approach evaluated the variability in the model’s outcomes due to the uncertainties in its input parameters, thereby identifying the critical parameters that are the most important drivers of uncertainty in the cost-effectiveness estimates. Key assumptions that varied in the analysis included time horizon (lifetime: up to 100 years versus 5 and 10 years), probabilities of all-cause mortality up to ± 6% and strategy-specific MACE according to no-overlap and maximum overlap between events assumptions estimates, cost estimates to reflect the 10th to 90th percentiles, and the discount rate between 0 to 5%. Further 2 or 3-way sensitivity analyses were performed to estimate the ranges of these individual parameter variations (Table 1).

**Table 1:**
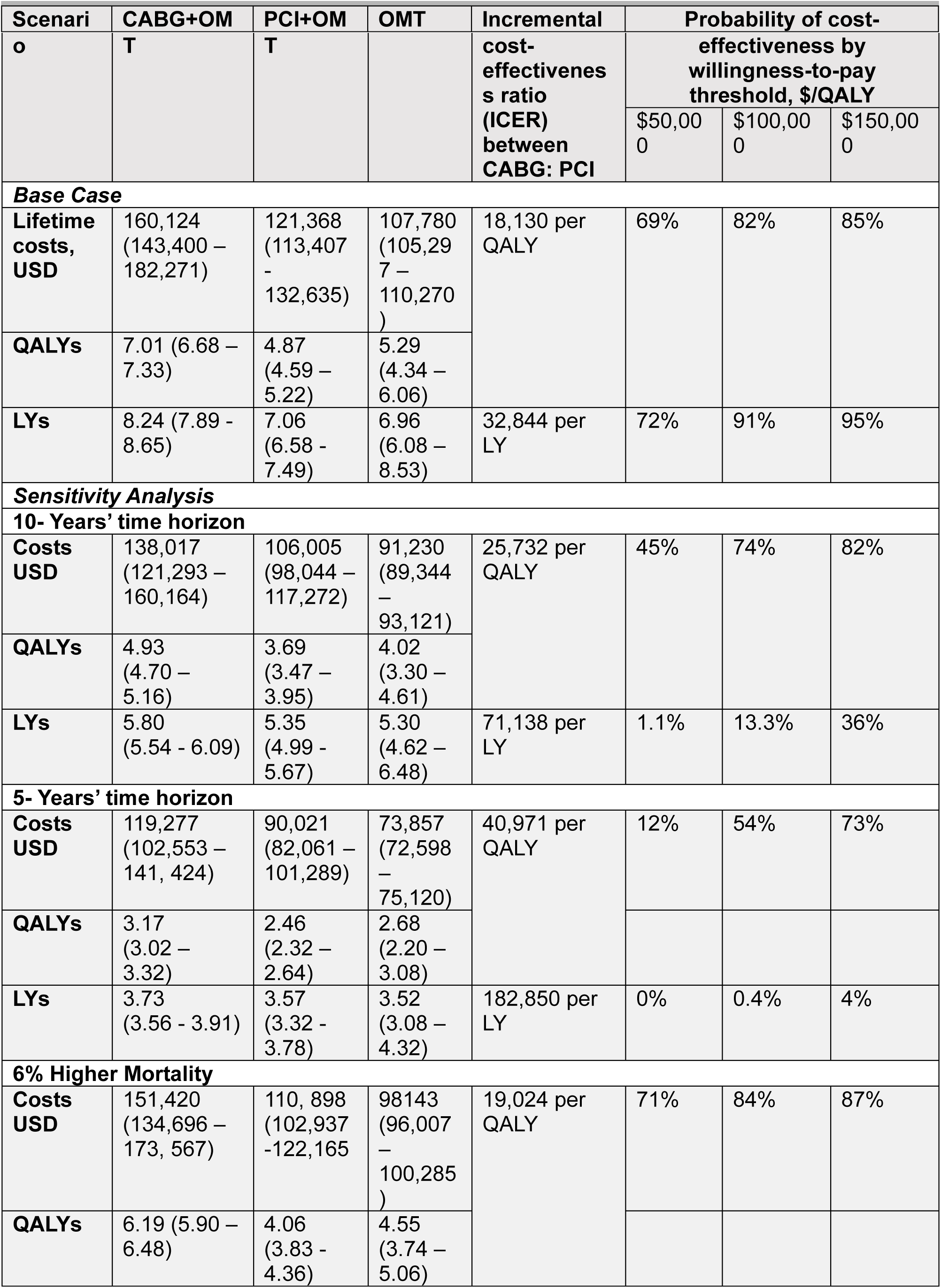
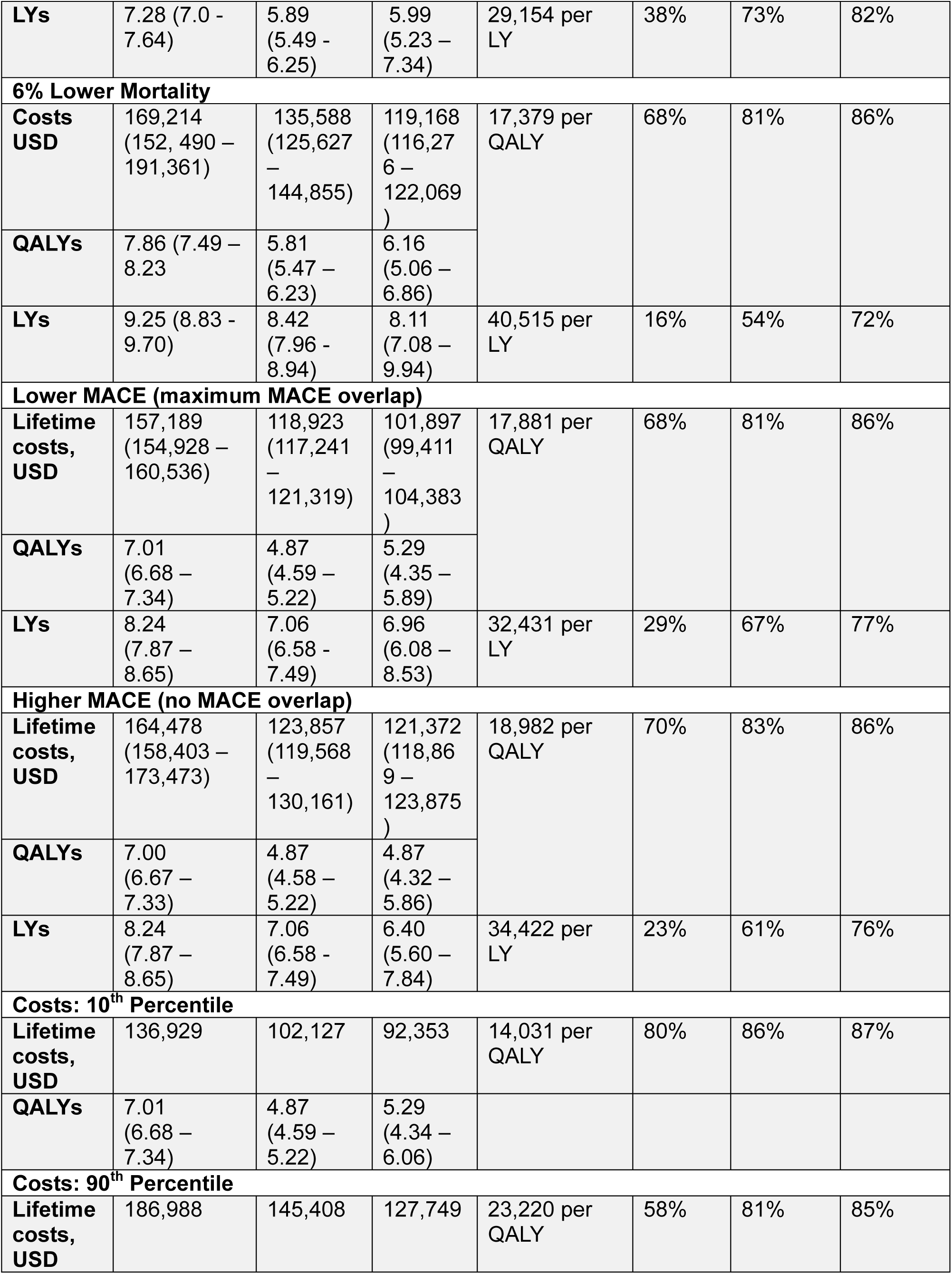
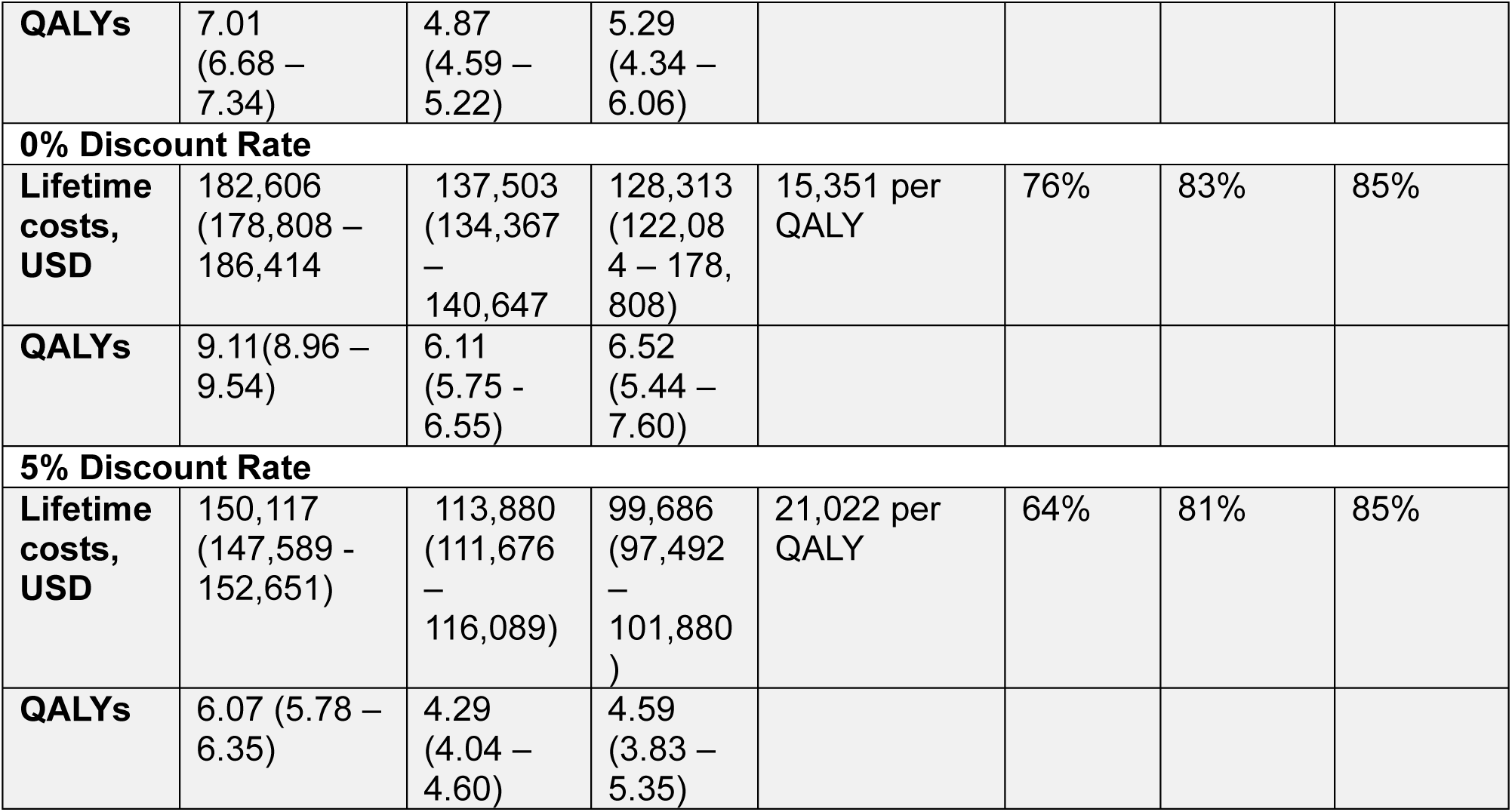
Base Case and Sensitivity Analysis: Discounted Cost, Life Expectancy and Cost-Effectiveness in STICH and REVIVED Trials.

We also performed a probabilistic sensitivity analysis; wherein multiple parameters were randomly varied simultaneously over predefined probability distributions via Monte Carlo simulations for 1000 iterations for each simulated cohort to determine the stability of our model. Beta distributions were applied to all probabilities and utilities, and gamma distributions to all costs. Distributions were fitted to mirror ranges examined in 1-way sensitivity analyses. Cost-effectiveness was displayed on the incremental cost-effectiveness plane and summarized with cost-effectiveness acceptability curves (Figure S2). As there is currently no established willingness-to-pay (WTP) threshold in US, WTP was set at $100,000/QALY gained, a commonly cited US benchmark^28^. We also considered the value taxonomy proposed by the American College of Cardiology and American Heart Association: high value represents either cost savings or an ICER <$50 000 per QALY gained; intermediate value is represented by ICERs between $50 000 and <$150 000 per QALY gained; and low value is described by ICERs ≥$150 000 per QALY gained^29^.

To assess the agreement of our model with the STICH and REVIVED trials, we compared the model’s estimates of undiscounted life expectancy and survival probabilities for the CABG+OMT strategy with the restricted mean survival times and mortality rates reported for the CABG group in the STICH trial. Additionally, we compared the model generated QALYs and survival probabilities for the PCI+OMT strategy against the predicted total QALYs and mortality percentages for PCI arm detailed in the cost-effectiveness analysis of the REVIVED trial.

Model parameter values are depicted in Table S1. Modeling was performed in TreeAge Pro Healthcare 2024 (TreeAge Software, Williamstown MA).

## 4. RESULTS

### Model Agreement

The estimated life expectancies for the CABG+OMT strategy align closely with the restricted mean survival time observed for the CABG group in the STICH trial over 5-years at 3.73 years (range 3.56 – 3.91) versus 3.94 years (range 3.80 – 4.08) as well as 10-years at 5.80 years (range 5.54 – 6.09) versus 6.51 years (range 6.21 – 6.80) respectively. Similarly, the model’s mortality estimates for CABG+OMT strategy mirrored the observed mortality rates for the CABG arm over the 9.8-year (or 117.6 months) duration of the STICH trial (Figure S2). Furthermore, the modeled QALYs for the PCI+OMT strategy at 10-years, 3.69 (range 3.47 - 3.95), approximate the predicted total QALYs for the PCI arm at 4.14 (range 4.02 – 4.27) over 8-years period. Additionally, the model’s estimated mortality for the PCI+OMT strategy was similar to the REVIVED trial’s reported mortality rates for the PCI arm at 41 months (Figure S3). The minor discrepancies in life expectancies may be attributable to the inclusion of MACE in survival probability calculations in the current analysis for CABG+OMT. Additionally, the differences in the duration of the follow-up periods— 8-years in the REVIVED trial versus 10-years in the current analysis—may explain the variance in projected total QALYs for the PCI+OMT group.

### Base Case Analysis (Lifetime Horizon)

After discounting and assuming sustained strategy specific event probabilities and utilities, the OMT strategy accrued a per person average of 5.29 QALYs or 6.96 LYs and a lifetime cost of $107,780, positioning it as the least costly strategy. The PCI+OMT strategy yielded 4.87 QALYs or 7.06 LYs and cost $121,368, or $13,588 per QALY gained relative to OMT, while the CABG+OMT strategy resulted in 7.01 QALYs or 8.24 LYs and cost $160,124 or $38,755 per QALY gained compared to PCI+OMT with an ICER of $18,130 per QALY gained. This places CABG+OMT strategy as high value according to American College of Cardiology and American Heart Association cost-effectiveness benchmarks for yielding an ICER <$50,000/QALY. Hence, CABG+OMT emerges as the preferred strategy at $50,000, $100,000 as well as $150,000 per QALY thresholds over lifetime horizon (Table 1). In probabilistic sensitivity analysis, CABG+OMT was the preferred strategy in 69% of iterations at a $50,000 threshold, in 82% of iterations at a $100,000 threshold, and in 85% of iterations at a $150,000 per QALY gained threshold (Figure 1 and 2).

**Figure 1:**
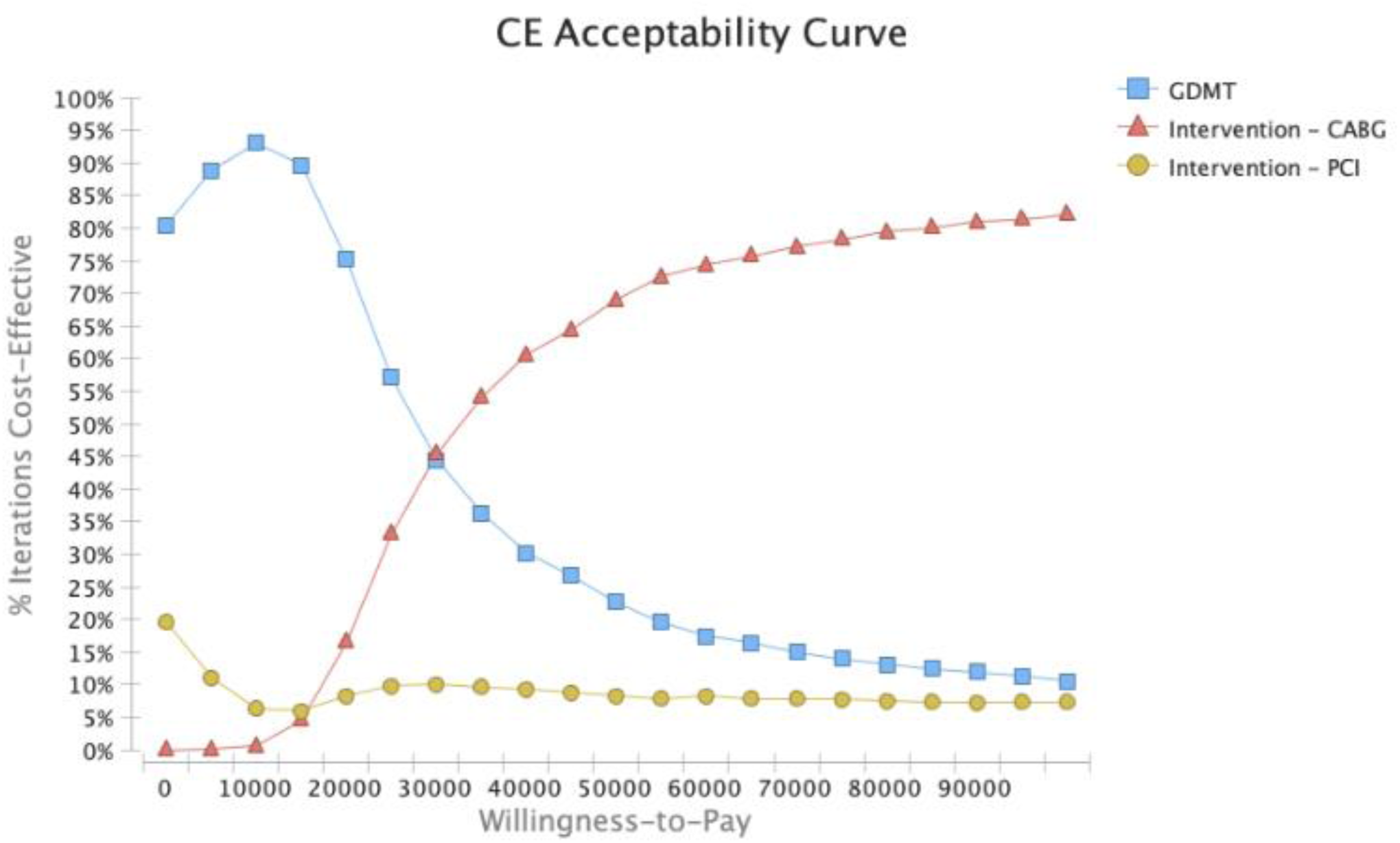
Cost Effectiveness (CE) acceptability curve shows the probability of CABG+OMT (red line with triangles) being cost-effective over a range of willingness-to-pay thresholds for severe ischemic cardiomyopathy cohort over lifetime horizon

**Figure 2:**
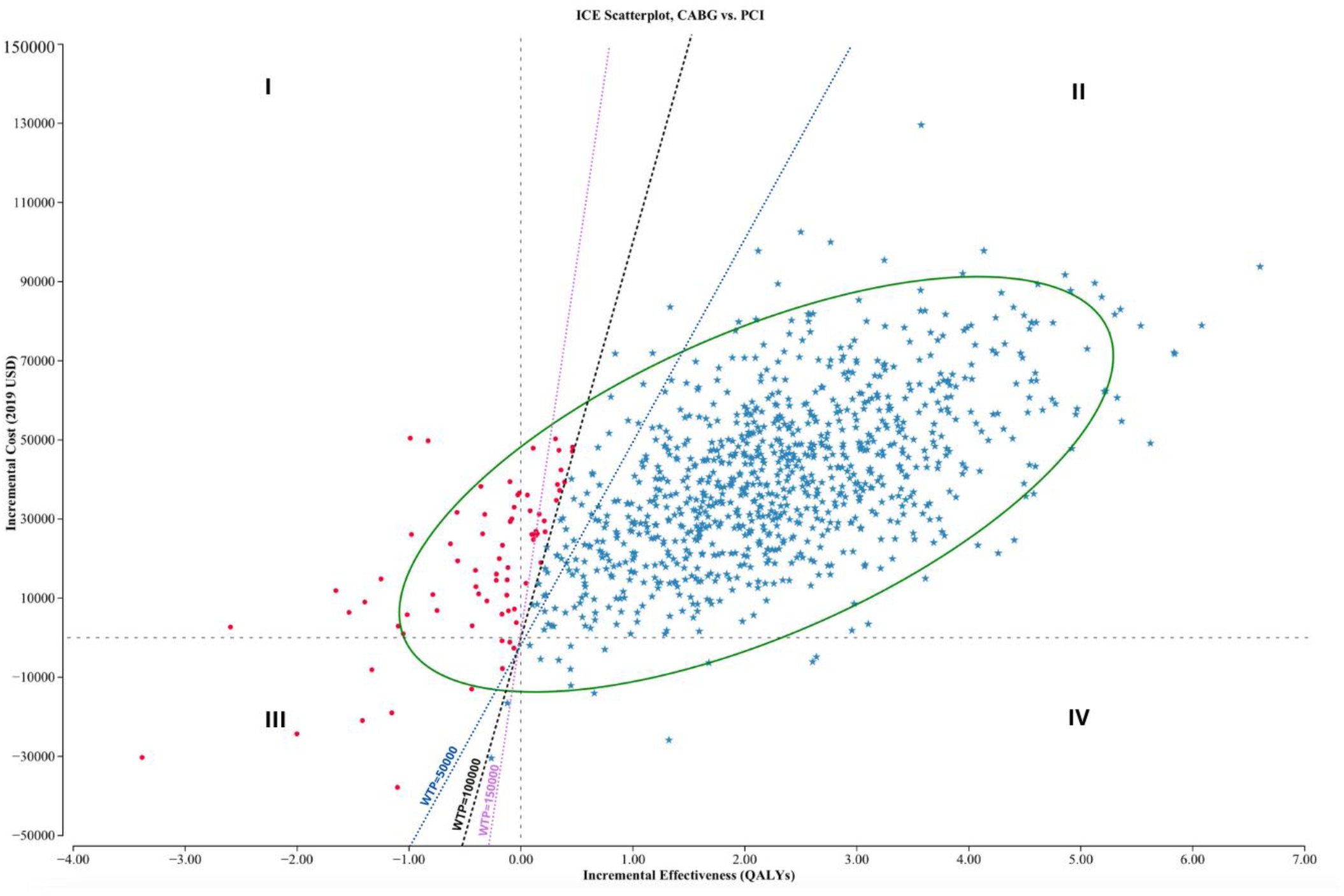
Incremental cost-effectiveness (ICE) scatterplot comparing coronary artery bypass grafting (CABG) plus optimal medical therapy (OMT) with percutaneous coronary intervention (PCI) plus optimal medical therapy (OMT). Quadrant I represent scenarios where CABG+OMT is more costly and less effective. Quadrant II represents scenarios where CABG+OMT is more costly and more effective. Quadrant III represents scenarios where CABG+OMT is less costly but more effective. Quadrant IV represents scenarios where CABG+OMT is less costly and less effective. QALY indicates quality-adjusted life-year; and WTP, willingness to pay.

### Sensitivity Analysis

The parameters with greatest influence over the comparative analysis between CABG+OMT and PCI+OMT are illustrated in the tornado diagram (Figure 3). The cost of CABG is the most impactful factor followed by the cost of PCI. Other important factors include the mortality risks associated with both CABG and PCI as well as their respective post-procedure utilities. 1-way deterministic sensitivity analyses of these parameters within the specified ranges does not result in a scenario where PCI would be favored over CABG. Furthermore, CABG+OMT strategy remains cost-effective at WTP thresholds of $50,000, $100,000, and $150,000 per QALY gained across all evaluated scenarios.

**Figure 3:**
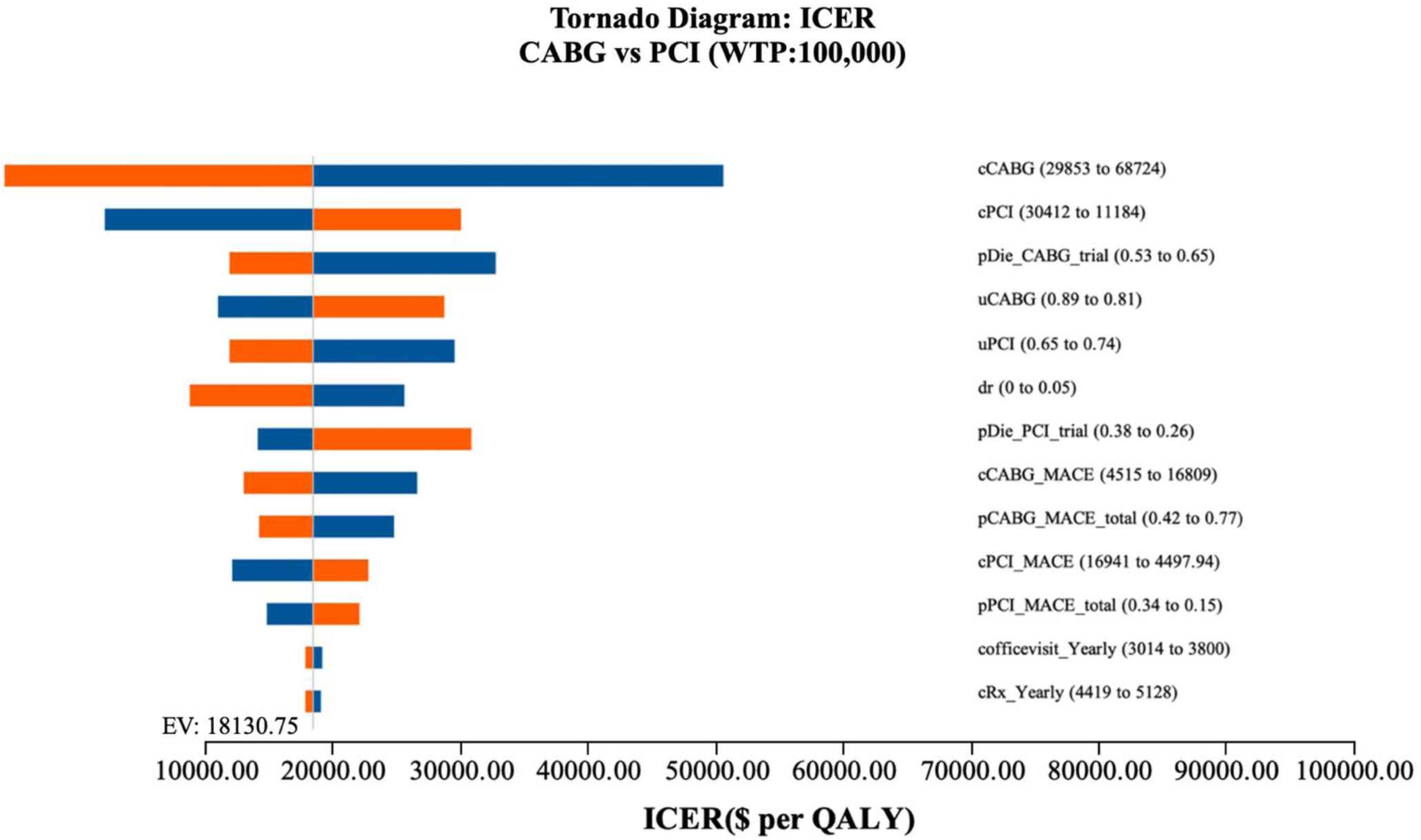
Tornado diagram summarizing 1-way sensitivity analyses on incremental cost-effectiveness ratio (cost per QALY gained). Blue and orange bars represent the effects of the upper and lower bounds of each input parameter, respectively.

### Projected costs and utilities over 5 and 10-years period

Within a 5-year timeframe, the OMT strategy accumulated an average of 2.68 QALYs or 3.52 LYs with a total cost of $73,857. By comparison, the PCI+OMT strategy achieved 2.46 QALYs or 3.57 LYs, costing $90,021. Concurrently, the CABG+OMT strategy yielded 3.17 QALYs or 3.73 LYs at an expense of $119,277, translating to $29,256 per QALY gained against PCI+OMT. The resulting ICER of $40,971 per QALY positions CABG+OMT as a valuable and economically beneficial choice, supported by probabilistic sensitivity analyses demonstrating favorability in 12%, 54%, and 73% of iterations at WTP thresholds of $50,000, $100,000, and $150,000 per QALY, respectively.

Over a 10-year period, the OMT strategy achieved an average of 4.02 QALYs or 5.30 LYs at a cost of $91,230, while PCI+OMT generated 3.69 QALYs at an increased cost of $106,005. The CABG+OMT strategy delivered 4.93 QALYs for $138,017, or $32,012 per QALY gained relative to PCI+OMT. This established an ICER of $25,732 per QALY, reaffirming CABG+OMT’s status as a high-value and economically advantageous option, as evidenced by 45%, 74%, and 82% of iterations favoring this strategy in probabilistic sensitivity analyses at the same WTP thresholds.

### Mortality and MACE probabilities variations

Varying strategy-specific mortalities by 6% alters their costs, QALYs, LYs and ICERs, nevertheless, CABG+OMT remains the most financially preferred strategy at $50,000, $100,000, and $150,000 per QALY WTP thresholds. Similarly, under assumptions of maximum and no MACE overlap potentially causing lower and higher MACE probabilities respectively, CABG+OMT strategy continues to dominate PCI+OMT and OMT across these WTP thresholds. Of note, the QALYs and LYs with higher or lower MACE probabilities are same as that over lifetime horizon because MACE is considered a temporary state in the analysis with assumption that upon resolution of MACE, patient return to chronic health state specific to each strategy.

### Costs and discount rates variations

Costs variations between 10^th^ and 90^th^ percentile and discount rates between 0-5% sustain CABG+OMT strategy economic dominance over PCI+OMT and OMT across WTP threshold spectrum. While QALYs are not impacted by costs variations, discount rates affect QALYs because utilities are discounted in addition to costs.

### Threshold Analysis

At $100,000 WTP threshold, CABG+OMT ceases to be the economically preferred strategy if the time horizon in less than 54 months, the mortality risk with CABG procedure surpasses 73% or the procedural cost associated with CABG strategy exceed $165,680.

### Is OMT cost-effective than PCI+OMT over lifetime horizon?

Although OMT alone is economically dominated by CABG, it appears to offer a substantial QALY gain of 5.29 at a cost of $107,780, rendering it relatively more cost-effective than PCI+OMT over a lifetime period. This is in contrast to the REVIVED trial results and its follow-up cost-effectiveness analysis where no clinical or economical difference was found between OMT and PCI+OMT strategies after 41 months^13,15^. An important factor that can explain this discrepancy is that OMT strategy was modeled based on the average of outcome estimates from STICH and REVIVED trials’ medical therapy only groups. STICH trial has reported better utility values in their medical therapy group compared to REVIVED over the trial’s duration. This could be since STICH has relatively younger participants than REVIVED. When STICH trial medical therapy group utility values are approximated to that of REVIVED in sensitivity analysis, OMT no longer retains its economic advantage over PCI+OMT at $100,000 per QALY WTP benchmark.

## 5. DISCUSSION

Our study shows that CABG as an adjunctive therapy to OMT represents the most cost-effective strategy over lifetime from a US healthcare perspective in patients with severe ICM, as compared to PCI+OMT or OMT only approaches. Over 5-year, 10-year and lifetime time horizons, CABG+OMT increases quality-adjusted life expectancy at a higher cost that is offset by lower risk of mortality and MACE relative to the competing strategies. While OMT is the least expensive option and PCI+OMT is less costly in our base case analysis, both are economically unfavorable to CABG+OMT, which yields an ICER of $30,531 per QALY gained compared to OMT alone and $18,130 per QALY gained compared to PCI+OMT. These values fall well below all commonly cited US WTP thresholds, categorizing CABG+OMT as a high-value and cost-effective approach in severe ICM according to the American College of Cardiology/American Heart Association cost and value framework.

This is the first cost-effectiveness study to investigate a head-to-head cost-utility comparison between CABG and PCI as a supplemental therapy to OMT in patients with severe ICM by using trial-level data, and decision-analysis modeling techniques to simulate long-term outcomes. In the absence of a clinical trial directly comparing these two interventions in this patient population, there exists an equipoise in the decision-making process regarding the optimal therapeutic approach. Our findings substantially contribute to filling this gap and are particularly crucial and timely as an increasing number of people are now living with ischemic cardiomyopathy as a result of early-onset ischemic heart disease through acquisition of cardiovascular risk factors (diabetes, hypertension, obesity etc.) and recent advances in interventional techniques reducing mortality from acute coronary syndromes^30^. ICM can progress to HFrEF, even with appropriate use of OMT if underlying ongoing ischemic injuries remain unaddressed^31^. The increasing prevalence of ischemia-induced HFrEF poses significant clinical and financial burdens for both patients and the healthcare system^32,33^. Identifying the most cost-effective strategy, CABG+ OMT in this case, that maximizes value for investment in such patients not only supports healthcare providers in making informed decisions to improve ischemic HFrEF outcomes but also aids policymakers in optimizing healthcare resource allocation.

Our study also benefits from extensive sensitivity analyses that test the robustness of model results to variations in input parameters to identify the most influential variables. The costs of index procedures (CABG and PCI) exerted the most influence followed by the probability of mortality associated with CABG and chronic health state utility values post-CABG and PCI. Variation of discount rates and mortality probabilities associated with PCI showed a moderate effect on model results, while the costs and probabilities of MACE for the CABG+OMT and PCI+OMT strategies, along with the costs of OMT alone, exerted minimal influence on results. Under various tested assumptions, the CABG+OMT strategy consistently emerged as the most cost-effective option. We also identified the extreme scenarios under which CABG+OMT strategy is not favorable at a threshold of $100,000 per QALY. These scenarios include an initial procedural cost exceeding $165,680, a risk of mortality from CABG surgery of 73% or higher, and a life expectancy of 54 months or less.

There is scarcity of data about cost-effectiveness analysis comparing CABG with PCI as complementary strategies to OMT in patients with severe stable ICM due to lack of large-scale clinical trials directly comparing the two strategies in this population and minimal inclusion or complete exclusion of such patients in prior studies^34–36^. Frequently, studies have focused on contrasting these interventions with medical therapy alone, thereby constraining the generalizability of findings to direct comparisons between CABG and PCI. Notable studies in this context include STICH trial and its follow-up cost-effectiveness analysis as well as REVIVED-BCIS2 trial and its follow-up cost-effectiveness analysis demonstrating that CABG and not PCI, used in conjunction with OMT is clinically and financially superior to OMT only approach^12–15^. These findings corroborate our current analysis, which reaffirms that the CABG+OMT strategy entails higher initial expenditures, predominantly due to substantial upfront costs associated with the CABG procedure in comparison to the PCI+OMT or OMT-only strategies in patients with severe ICM and an EF of 35% or less. Nonetheless, these increased costs are offset by significant reductions in MACE, prolongation of life expectancy, and improvements in QoL measures, thereby establishing CABG+OMT as the most cost-effective option. Additionally, our sensitivity analyses identify key factors that contribute to the economic attractiveness of the CABG+OMT strategy. These include the costs associated with the CABG and PCI procedures, the mortality risks linked to these interventions, and the utility values associated with these procedures.

### Limitations

Our study also has several limitations. First, significant differences in baseline characteristics between the STICH and REVIVED trials could impact analysis estimates. For instance, the STICH cohort was relatively younger with a mean age of 60 years, compared to the REVIVED cohort’s mean age of 69 years. Moreover, the use of OMT and preventive device therapies was more prevalent in REVIVED than in STICH. These variations could influence the mortality and morbidity risks in these trials. We addressed these disparities by conducting sensitivity analyses and varying mortality and MACE risks across a wide range to provide estimates under various assumptions. All the tested assumptions favored CABG over PCI in our analysis.

An important caveat to be considered while interpreting the results of current analysis is that we used probabilities instead of hazard ratios to model mortality and MACE due to lack of access to patient level data. While probabilities provide time-independent estimates of outcomes, they may underestimate time-varying risks such as increased procedure-related adverse events risks in the immediate post-procedure period. Therefore, the applicability of the study results to scenarios with increased procedural risks is constrained. Furthermore, MACE probabilities were estimated based on assumptions of maximum and no overlap between adverse events, which could under- or overestimate the true MACE risks. Our sensitivity analysis offered estimates under both assumptions; however, the actual estimates likely reside between these extremes. Similarly, MACE was modeled as a transient state in this analysis with the anticipation that patients would return to their baseline chronic health state following the resolution of the event. However, this assumption may not hold true in real-world, where individuals could experience a permanent decrease in health utility following MACE. Consequently, in actual clinical settings, patients may not regain their pre-MACE level of health utility, resulting in a chronic health state that is diminished compared to prior conditions.

Additionally, this cost-effectiveness analysis was performed from a US healthcare perspective; therefore, results of current analysis can have different estimates in other economic healthcare systems. Also, current analysis does not account for variations in the level of care during hospitalizations for MACE, such as intensive care versus standard medical ward care. Therefore, should a higher level of care be predominant during hospitalization, the associated costs with that strategy could increase significantly.

Notably, STICH did not assess bleeding risks, and REVIVED did not include stroke risks; consequently, these events were not incorporated into the MACE calculations. This omission limits the applicability of this analysis for individuals at heightened risk for these conditions after index procedures.

Lastly, this analysis lacks a sensitivity analysis based on various ejection fraction (EF) levels and the number and types of coronary arteries involved to assess the magnitude of differential advantage of CABG over PCI at certain EF thresholds as well as degree of ischemia. This is due to the REVIVED trial’s lack of analysis on the individual components of its composite primary outcome for these specific parameters.

## 6. CONCLUSION

CABG+OMT is the most cost-effective strategy in patients with severe ICM as compared with PCI+OMT or OMT only strategies but is sensitive to procedural costs, mortality risks and QoL utilities associated with CABG and PCI. Further studies are warranted to validate these findings and optimize individualized treatment decisions.

## Data Availability

The corresponding and senior authors have full access to all the data in the study and take responsibility for the integrity of the data and the accuracy of the data analysis.

## 7. ACKNOWLEDGMENTS

We express our gratitude to Dr. Samir Saba, MD, and Dr. Andrew Lawrence Wickerham, MD, MPH, MBA, for their valuable and insightful feedback and thorough review of this study.

## 8. DISCLOSURE STATEMENT

The authors declare no conflicts of interest related to this study. All authors have reviewed and approved the final manuscript for submission. The corresponding and senior authors have full access to all the data in the study and take responsibility for the integrity of the data and the accuracy of the data analysis.

## 9. SOURCES OF FUNDING

This study did not receive any funding.

## Notes

### Competing Interest Statement

The authors have declared no competing interest.

### Funding Statement

The study did not receive any funding.

